# Predicting the effect of statins on cancer risk using genetic variants: a Mendelian randomization study in UK Biobank

**DOI:** 10.1101/2020.02.28.20028902

**Authors:** Paul Carter, Mathew Vithayathil, Siddhartha Kar, Rahul Potluri, Amy M. Mason, Susanna C. Larsson, Stephen Burgess

**Affiliations:** Department of Public Health and Primary Care, University of Cambridge, Cambridge, UK; MRC Cancer Unit, University of Cambridge, Cambridge, UK; MRC Integrative Epidemiology Unit, University of Bristol, Bristol, UK; Population Health Sciences, Bristol Medical School, University of Bristol, Bristol, UK; Unit of Cardiovascular and Nutritional Epidemiology, Institute of Environmental Medicine, Karolinska Institutet, Stockholm, Sweden; Department of Surgical Sciences, Uppsala University, Uppsala, Sweden; MRC Biostatistics Unit, University of Cambridge, Cambridge, UK

**Keywords:** Mendelian randomization, cancer risk, lipids, statins, genetic epidemiology

## Abstract

Laboratory studies have suggested oncogenic roles of lipids, as well as anticarcinogenic effects of statins. We here assess the potential effect of statin therapy on cancer risk in Mendelian randomization analyses. We obtained genetic associations with the risk of overall and 22 site-specific cancers for 367,703 individuals in UK Biobank. In total, 75,037 individuals had a cancer event. Variants in the *HMGCR* gene region, which represent proxies for statin treatment, were associated with overall cancer risk (OR per 1 standard deviation increase in LDL-cholesterol 1.32, 95% CI 1.13-1.53, p=0.0003), but variants in gene regions representing alternative lipid-lowering treatment targets (*PCSK9, LDLR, NPC1L1, APOC3, LPL*) were not. Genetically-predicted LDL-cholesterol was not associated with overall cancer risk (OR 1.01, 95% CI 0.98-1.05, p=0.50). Our results predict that statins reduce cancer risk, but other lipid-lowering treatments do not. This suggests that statins reduce cancer risk through a cholesterol independent pathway.

---

Statins are inhibitors of 3-hydroxy-3-methyl-glutaryl-coenzyme A reductase (HMGCR), which is the rate-limiting enzyme in the melavonate pathway; a pathway producing a range of cell signalling molecules with the potential to regulate oncogenesis. This is supported by strong laboratory evidence that statins induce anticarcinogenic effects on cell proliferation and survival in various cell lines^1–4^, and reduce tumour growth in a range of *in vivo* models^5–10^. Furthermore, epidemiological studies of pre-diagnostic use of statins have been associated with reduced risk of specific cancer types^11–13^. However, meta-analyses of cardiovascular-focused randomized controlled trials have shown no effect of statins on cancer^14,15^. Conclusions from these trials are limited as they lack adequate power and longitudinal follow-up necessary for assessing impact on cancer risk. At present, no clinical trials have been designed to assess the role of statins in primary cancer prevention and their role in chemoprevention remains uncertain.

A putative protective effect of statins on cancer development could be through either cholesterol dependent or independent effects^16–19^. Cholesterol is a key mediator produced by the mevalonate pathway and is essential to cell signalling and membrane structure, with evidence demonstrating the potential to drive oncogenic processes and tumour growth^20,21^. However, the epidemiological relationships between circulating cholesterol and cancer risk remain unclear. Individual observational studies have reported positive^22,23^, inverse^22–25^ and no association^26–29^ between circulating levels of total cholesterol, low-density lipoprotein (LDL) cholesterol, high-density lipoprotein (HDL) cholesterol, and triglycerides with risk of overall and site-specific cancers. Different cancer types have distinct underlying pathophysiology, and meta-analyses of observational studies highlight a likely complex relationship which varies according to both lipid fraction^30,31^ and cancer type ^29,32–34^. Furthermore, cancer can lower cholesterol levels for up to 20 months prior to diagnosis^35^. Thus, the true relationship between lipids and cancer development remains equivocal.

Mendelian randomization is an epidemiological approach which assesses associations between genetically-predicted levels of a risk factor and a disease outcome in order to predict the causal effect of the risk factor on an outcome^36^. The use of genetic variants minimizes the influences of reverse causality and confounding factors on estimates. Mendelian randomization studies also have the potential to predict the outcomes of trials for specific therapeutic interventions. A limited number of Mendelian randomization studies have investigated the relationship between HMGCR inhibition and cancer^37–41^, with protective associations observed for prostate cancer^37^, colorectal cancer^38^, breast cancer^39,40^, and ovarian cancer^41^. However, no comprehensive Mendelian randomization investigation has evaluated the predicted impact of HMGCR inhibition or the causal role of specific lipid fractions on the risk of many of the most common site-specific cancers.

We here investigate the relationship between HMGCR inhibition and the risk of overall cancer and site-specific cancers using genetic variants in the *HMGCR* gene region. To understand whether statins may influence cancer risk through lipid-related mechanisms, we also assess the relationship between lipids and cancer risk by polygenic Mendelian randomization analyses using common lipid-associated genetic variants. Additionally, to mimic other lipid-lowering pharmaceutical interventions, gene-specific analyses were performed using variants in or near gene regions targeted by these therapies.

Baseline characteristics of the participants in the UK Biobank and numbers of outcomes are provided in Table 1. In total, 75,037 of the participants had a cancer event, of which 48,674 participants had one of the 22 defined site-specific cancers. Power calculations for the various analyses are presented in Supplementary Figure 1 (site-specific cancers) and Supplementary Table 1 (overall cancer). Gene-specific analyses were only well-powered for overall cancer. Polygenic analyses were well-powered to detect moderate effects for overall cancer and for common site-specific cancers, but less well-powered for less common site-specific cancers.

**Table 1:** Baseline characteristics of UK Biobank participants included in this study and numbers of outcome events

| Characteristic or cancer site/type | Mean (SD) or N (%)† |
| --- | --- |
| Sample size | 367,703 (100) |
| Female | 198,904 (54.1) |
| Age at baseline | 57.2 (8.1) |
| Body mass index | 27.3 (4.8) |
| Systolic blood pressure | 137.6 (18.6) |
| Diastolic blood pressure | 82.0 (10.1) |
| Smoking status (current / ex / never)* | 37,866 (10.3) / 185,704 (50.5) / 143,777 (39.1) |
| Alcohol status (current / ex / never)* | 342,797 (93.2) / 12,732 (3.5) / 11,646 (3.2) |
| History of type 2 diabetes | 15,834 (4.3) |
| Overall cancer | 75,037 (20.4) |
| Breast | 13,666 (6.9) |
| Prostate | 7872 (4.7) |
| Lung | 2838 (0.8) |
| Bowel | 5486 (1.5) |
| Melanoma | 4869 (1.3) |
| Non-Hodgkin's lymphoma | 2296 (0.6) |
| Kidney | 1310 (0.4) |
| Head/Neck | 1615 (0.4) |
| Brain | 810 (0.2) |
| Bladder | 2588 (0.7) |
| Pancreas | 1264 (0.3) |
| Uterus | 1931 (1.0) |
| Leukaemia | 1403 (0.4) |
| Oesophagus | 843 (0.2) |
| Ovaries | 1520 (0.8) |
| Gastric | 736 (0.2) |
| Liver | 324 (0.1) |
| Myeloma | 656 (0.2) |
| Thyroid | 375 (0.1) |
| Biliary | 387 (0.1) |
| Cervix | 1928 (1.0) |
| Testes | 735 (0.4) |

\* Excluding 356 participants with smoking status absent and 528 participants with alcohol consumption status absent
† For sex-specific cancers, this is the percentage of individuals of the relevant sex

Associations for specific gene regions representing targets of lipid-lowering drugs are displayed in Figure 1 and Supplementary Figures 2-7. For overall cancer, there was evidence of association for variants in the *HMGCR* gene region (OR 1.32, 95% CI 1.13-1.53, p=0.0003), but not for other gene regions: for *PCSK9* (OR 1.03, 95% CI 0.92-1.14, p=0.66), for *LDLR* (OR 0.99, 95% CI 0.92-1.07, p=0.86), for *NPC1L1* (OR 0.87, 95% CI 0.73-1.04, p=0.13), for *APOC3* (OR 1.08, 95% CI 0.98-1.19, p=0.15), or for *LPL* (OR 1.03, 95% CI 0.95-1.13, p=0.45). The association of variants in the *HMGCR* gene region with overall cancer remained broadly similar when restricting outcomes to the 48,674 individuals who had one of the 22 site-specific cancers (OR 1.29, 95% CI 1.08-1.54, p=0.005) and when excluding outcomes that were self-reported only (70,734 remaining cases, OR 1.30, 95% CI 1.12-1.52, p=0.0007).

**Figure 1:**
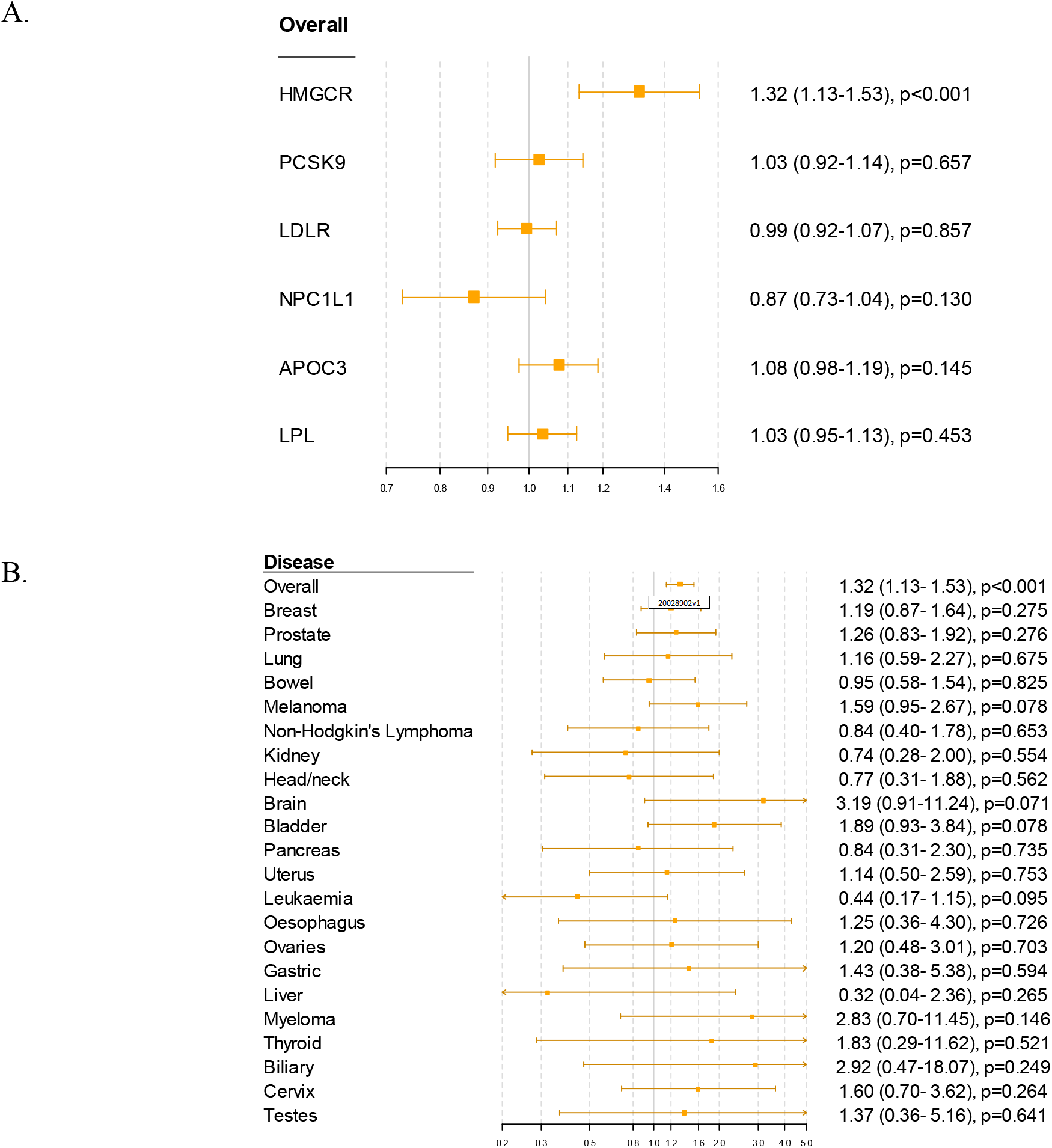
Gene-specific Mendelian randomization estimates (odds ratio with 95% confidence interval per 1 standard deviation increase in lipid fraction) for variants in gene regions representing targets of lipid-lowering treatments. Estimates are scaled to a 1 standard deviation increase in LDL-cholesterol for the *HMGCR, PCSK9, LDLR* and *NPC1L1* regions, and to a 1 standard deviation increase in triglycerides for the *APOC3* and *LPL* regions. A: associations with overall cancer for each gene region in turn. B: associations with site-specific cancers for variants in the *HMGCR* gene region.

For site-specific cancers, the *HMGCR* gene region showed positive associations for five of the six most common cancer sites (breast, prostate, melanoma, lung, and bladder; not for bowel), although none of these results individually reached a conventional level of statistical significance. There was little evidence for associations in site-specific analyses for other lipid-lowering drug targets.

Polygenic Mendelian randomization estimates are displayed in Figure 2 for HDL-cholesterol, LDL-cholesterol, and triglycerides (see also Supplementary Table 2), and Figure 3 for total cholesterol. For overall cancer, the odds ratio (OR) per 1 standard deviation increase in genetically-predicted levels of the risk factor was 1.01 (95% confidence interval [CI] 0.98-1.05, p=0.50) for LDL-cholesterol, 0.99 (95% CI 0.95-1.03, p=0.54) for HDL-cholesterol, 1.00 (95% CI 0.98-1.05, p=0.85) for triglycerides, and 1.01 (95% CI 0.98-1.05; p=0.57) for total cholesterol.

**Figure 2:**
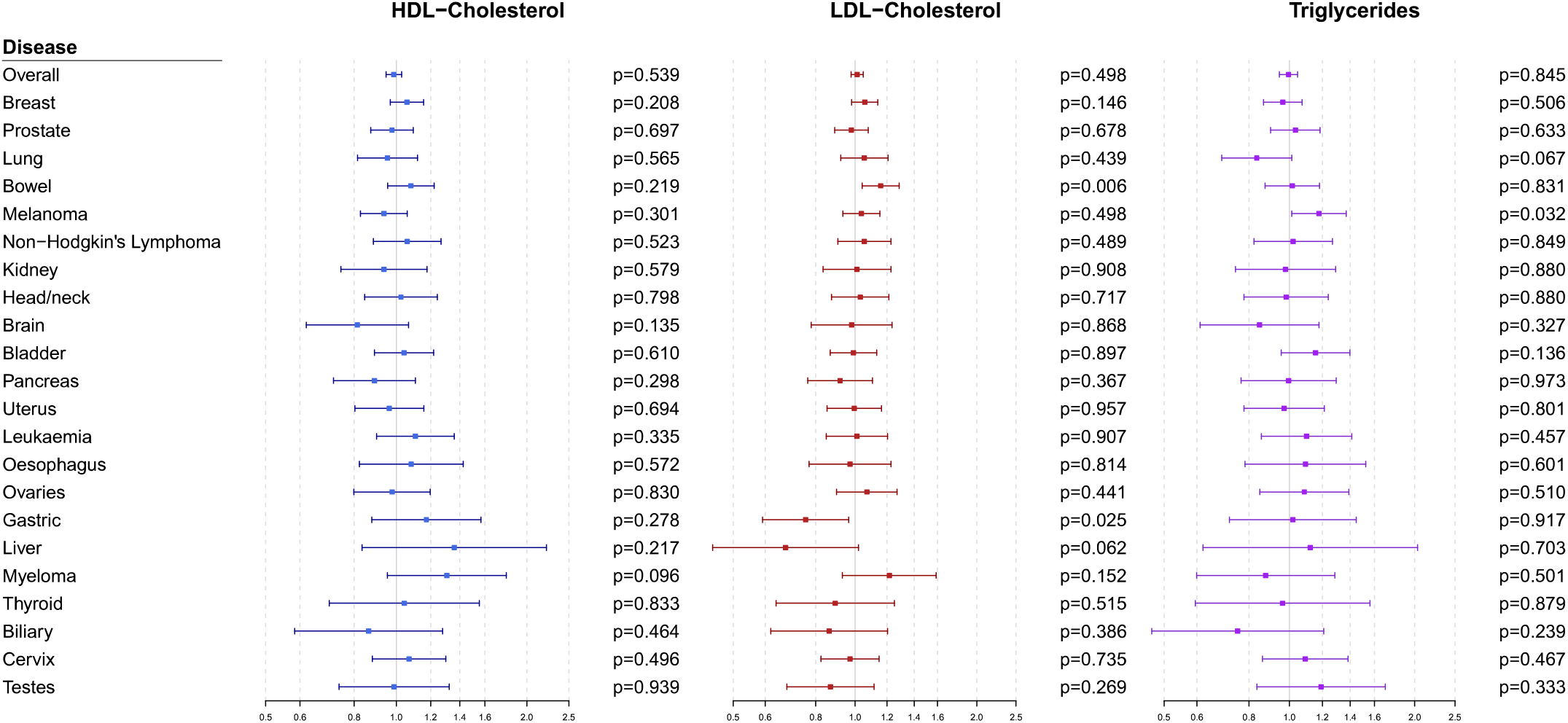
Multivariable Mendelian randomization estimates for HDL-cholesterol, LDL-cholesterol, and triglycerides (odds ratio with 95% confidence interval per 1 standard deviation increase in lipid fraction) from polygenic analyses including all lipid-associated variants

**Figure 3:**
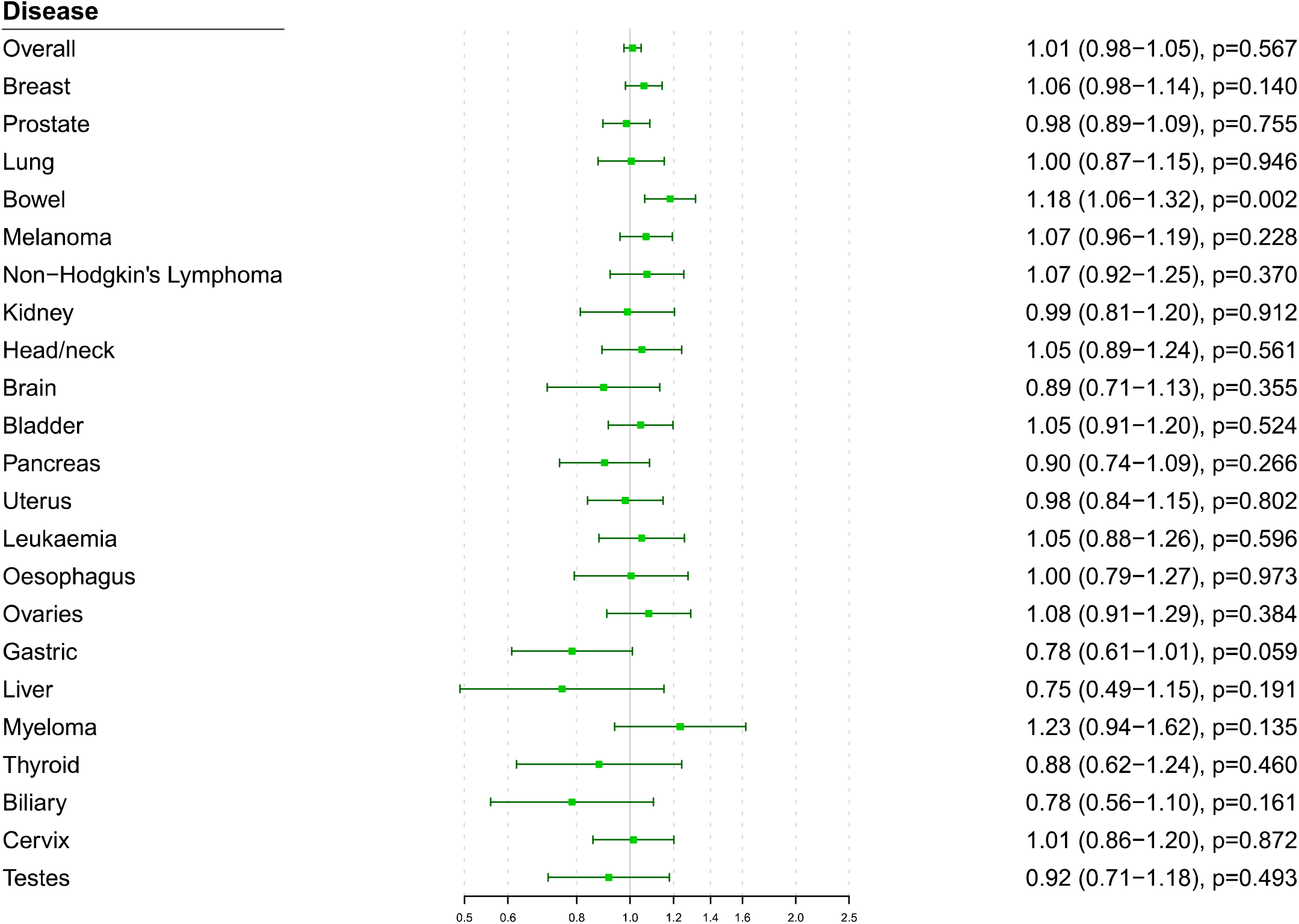
Univariable Mendelian randomization estimates for total cholesterol (odds ratio with 95% confidence interval per 1 standard deviation increase in lipid fraction) from polygenic analyses including all lipid-associated variants

For site-specific cancers, there were positive associations between risk of bowel cancer and genetically-predicted levels of total cholesterol (OR 1.18, 95% CI 1.06-1.32, p=0.002) and LDL-cholesterol (OR 1.16, CI 1.04-1.29, p=0.006). Results were attenuated in robust methods (Supplementary Table 3). No other associations were statistically significant after accounting for multiple testing.

Our comprehensive Mendelian randomization investigation shows a positive association between overall cancer and variants in the *HMGCR* gene region which can be considered as proxies for statin therapy. However, gene regions which can be considered as proxies for alternative lipid-lowering therapies were not associated with cancer risk. Furthermore, there was little consistent evidence of an association between genetically-predicted lipid fractions and cancer outcomes in polygenic analyses either for overall cancer or for any site-specific cancer. Taken together, our findings predict that statins lower the risk of cancer, and provide important mechanistic evidence that this occurs through mechanisms other than lipid lowering.

We found that genetic variants in the *HMGCR* region, serving as proxies for targets of statin therapy, were associated with a 26% decrease in risk of overall cancer per standard deviation (around 39 mg/dL or 1.0 mmol/L) reduction in genetically-predicted LDL-cholesterol. For coronary artery disease, the short-term impact of statins in trials is around one-third of the Mendelian randomization estimate (which represents the impact of lifelong reduced exposure)^42^. This suggests that any reduction in cancer risk from statins is likely to be modest. While our results should be seen as tentative until trials have demonstrated benefit, associations of *HMGCR* variants show broad concordance with statin therapy for many continuous phenotypes^43^, and suggest that statins reduce risk of coronary artery disease^44^, increase risk of Type 2 diabetes^45^, and increase risk of intracerebral haemorrhage^46,47^, as confirmed in clinical trials^48–50^.

The notion that statins could be used for chemoprevention is longstanding. Nobel Prize winners Goldstein and Brown proposed that this occurs through non-lipid lowering mechanisms^16^. We provide mechanistic evidence using human genetics supporting this theory. Our results suggest that with respect to genetically predicted HMGCR inhibition and cancer risk, LDL-cholesterol is simply a biomarker of HMGCR inhibition that is accessible, but the true causal pathway is likely via another molecule whose levels are correlated with its LDL-cholesterol lowering effect. HMGCR catalyses the rate-limiting step of the mevalonate pathway; a pathway with an arm leading to the end point of cholesterol synthesis and another arm leading to isoprenoid synthesis. Measuring levels of intra-cellular isoprenoids is challenging but these molecules are implicated in cancer via their role as major post-translational modifiers of key oncogenic proteins^17^. In particular, mevalonate and other isoprenoid metabolites are required for the prenylation and functioning of the Ras and Rho GTPases, which are oncoproteins, and involved in important cellular processes including apoptosis, phagocytosis, vascular trafficking, cell proliferation, transmigration, cytoskeleton organisation, and recruitment of inflammatory cells. Statin inhibition of these metabolites has demonstrated anti-oncological effects *in vivo* and *in vitro*^18^ including the promotion of tumour cell death and apoptosis^51–54^, inhibition of angiogenesis^55^, and reduction of tumour cell invasion and metastasis^56,57^. Other potential statin-mediated mechanisms of tumour suppression include the reduction of systemic inflammatory mediators like interleukin 1-beta and tumour necrosis factor^55,58^, and epigenetic regulation through inhibiting HMGCR-mediated deacetylation^59^ which contributes to colorectal cancer in mouse models^60^. Thus, our findings based on large-scale human genetic data are consistent with pre-clinical studies on statins in cancer which have repeatedly argued for a cholesterol independent mechanism for statin effects on cancer.

Our investigation has many strengths, but also limitations. The large sample size of over 360,000 participants and the broad set of outcomes analysed render this the most comprehensive Mendelian randomization analysis of lipids and cancer outcomes conducted to date. However, the investigation has a number of limitations. For many site-specific cancers, there were not enough outcome events to obtain adequate power to rule out the possibility of moderate causal effects. While there is evidence to support our assumption that genetic variants in relevant gene regions can be used as proxies for pharmacological interventions, our findings should be considered with caution until they have been replicated in clinical trials. Our investigation was able to compare subgroups of the population with different lifelong average levels of lipid fractions, but the impact of lowering a particular lipid fraction in practice is likely to differ from the genetic association, particularly quantitatively^61^. Finally, analyses were conducted in UK-based participants of European ancestries. While it is recommended to have a well-mixed study population for Mendelian randomization in order to ensure that genetic associations are not influenced by population stratification, it means that results may not be generalizable to other ethnicities or nationalities.

In conclusion, our findings suggest that HMGCR inhibition may have a chemopreventive role in cancer though non-lipid lowering properties, and that this role may apply across cancer sites. The efficacy of statins for cancer prevention must be urgently evaluated.

## ONLINE METHODS

### Study design and data sources

We performed two-sample Mendelian randomization analyses, taking genetic associations with risk factors (i.e., serum lipid levels) from one dataset, and genetic associations with cancer outcomes from an independent dataset, as performed previously for cardiovascular diseases^62^.

We obtained genetic associations with serum lipid concentrations (total cholesterol, LDL-cholesterol, HDL-cholesterol, and triglycerides) from the Global Lipids Genetic Consortium (GLGC) on up to 188,577 individuals of European ancestry^63^. Genetic associations were estimated with adjustment for age, sex, and genomic principal components within each participating study after inverse rank quantile normalization of lipid concentrations, and then meta-analysed across studies.

We estimated genetic associations with cancer outcomes on 367,703 unrelated individuals of European ancestry from UK Biobank, a population-based cohort recruited between 2006-2010 at 22 assessment centres throughout the UK and followed-up until 31st March 2017 or their date of death (recorded until 14th February 2018)^64^. We defined cancer outcomes for overall cancer and for the 22 most common site-specific cancers in the UK (Supplementary Table 4). Outcomes were based on electronic health records, hospital episode statistics data, national cancer registry data, and death certification data, which were all coded according to ICD-9 and ICD-10 diagnoses. Further cancer outcomes were captured by self-reported information validated by interview with a trained nurse, and from cancer histology data in the national cancer registry. To obtain genetic association estimates for each outcome, we conducted logistic regression with adjustment for age, sex, and 10 genomic principal components using the *snptest* software program. For sex-specific cancers (breast, uterus, and cervix for women, prostate and testes for men), analyses were restricted to individuals of the relevant sex.

### Gene-specific analyses for HMGCR and other drug proxy variants

We performed targeted analyses for variants in the *HMGCR* gene region that can be considered as proxies for statin therapy. Additionally, we conducted separate analyses for the *PCSK9, LDLR, NPC1L1, APOC3*, and *LPL* gene regions, mimicking other lipid altering therapies (Supplementary Table 5). These regions were chosen as they contain variants that explain enough variance in lipids to perform adequately powered analyses. Variants in each gene region explained 0.4% (*HMGCR*), 1.2% (*PCSK9*), 1.0% (*LDLR*), 0.2% (*NPC1L1*), 0.1% (*APOC3*) and <0.1% (*LPL*) of the variance in LDL-cholesterol. The *APOC3* and *LPL* variants also explained 1.0% and 0.9% of the variance in triglycerides respectively. We performed the inverse-variance weighted method accounting for correlations between the variants^65^.

Estimates for the *HMGCR, PCSK9, LDLR* and *NPC1L1* gene regions are scaled to a 1 standard deviation increase in LDL-cholesterol, whereas estimates for the *APOC3* and *LPL* gene regions are scaled to a 1 standard deviation increase in triglycerides.

### Polygenic analyses for all lipid-related variants

We carried out polygenic analyses based on 184 genetic variants previously demonstrated to be associated with at least one of total cholesterol, LDL-cholesterol, HDL-cholesterol or triglycerides at a genome-wide level of significance (*p* < 5 × 10^−8^) in the GLGC^66^. These variants explained 15.0% of the variance in total cholesterol, 14.6% in LDL-cholesterol, 13.7% in HDL-cholesterol, and 11.7% in triglycerides in the GLGC.

To obtain the associations of genetically-predicted values of LDL-cholesterol, HDL-cholesterol and triglycerides with each cancer outcome while accounting for measured genetic pleiotropy via each other, we performed multivariable Mendelian randomization analyses using the inverse-variance weighted method^67^. For total cholesterol, we performed univariable Mendelian randomization analyses using the inverse-variance weighted method^68^. To account for between-variant heterogeneity, we used random-effects models in all analyses. For polygenic analyses that provided evidence of a causal effect, we additionally performed robust methods for Mendelian randomization, in particular the MR-Egger^69^ and weighted median methods^70^. All estimates are expressed per 1 standard deviation increase in the corresponding lipid fraction (in the GLGC, 1 standard deviation was 45.6 mg/dL for total cholesterol, 39.0 mg/dL for LDL-cholesterol, 15.8 mg/dL for HDL-cholesterol, and 90.5 mg/dL for triglycerides).

As power calculators have not been developed for multivariable Mendelian randomization analyses, we performed power calculations for polygenic analyses based on univariable Mendelian randomization for each lipid fraction in turn, and for gene-specific analyses for each gene region in turn^71^. We carried out all analyses using R (version 3.4.4) unless otherwise stated. All statistical tests and p-values presented are two-sided.

## Data Availability

Data used in the manuscript is publicly available or else available to any bona fide researcher on application.

http://lipidgenetics.org/#data-downloads-title

https://biobank.ctsu.ox.ac.uk/crystal/

## Acknowledgements

Stephen Burgess is supported by Sir Henry Dale Fellowship jointly funded by the Wellcome Trust and the Royal Society (grant number 204623/Z/16/Z). Stephen Burgess and Amy Mason are supported by the UK National Institute for Health Research Cambridge Biomedical Research Centre.

## Author contributors

All authors were involved in study design, conduct of the study, analysis of data, interpretation of results, or re-drafting of this report. SB had full access to the combined data, performed the statistical analysis, and had responsibility for submission of the report for publication.

---

**Supplementary Figure 1:**
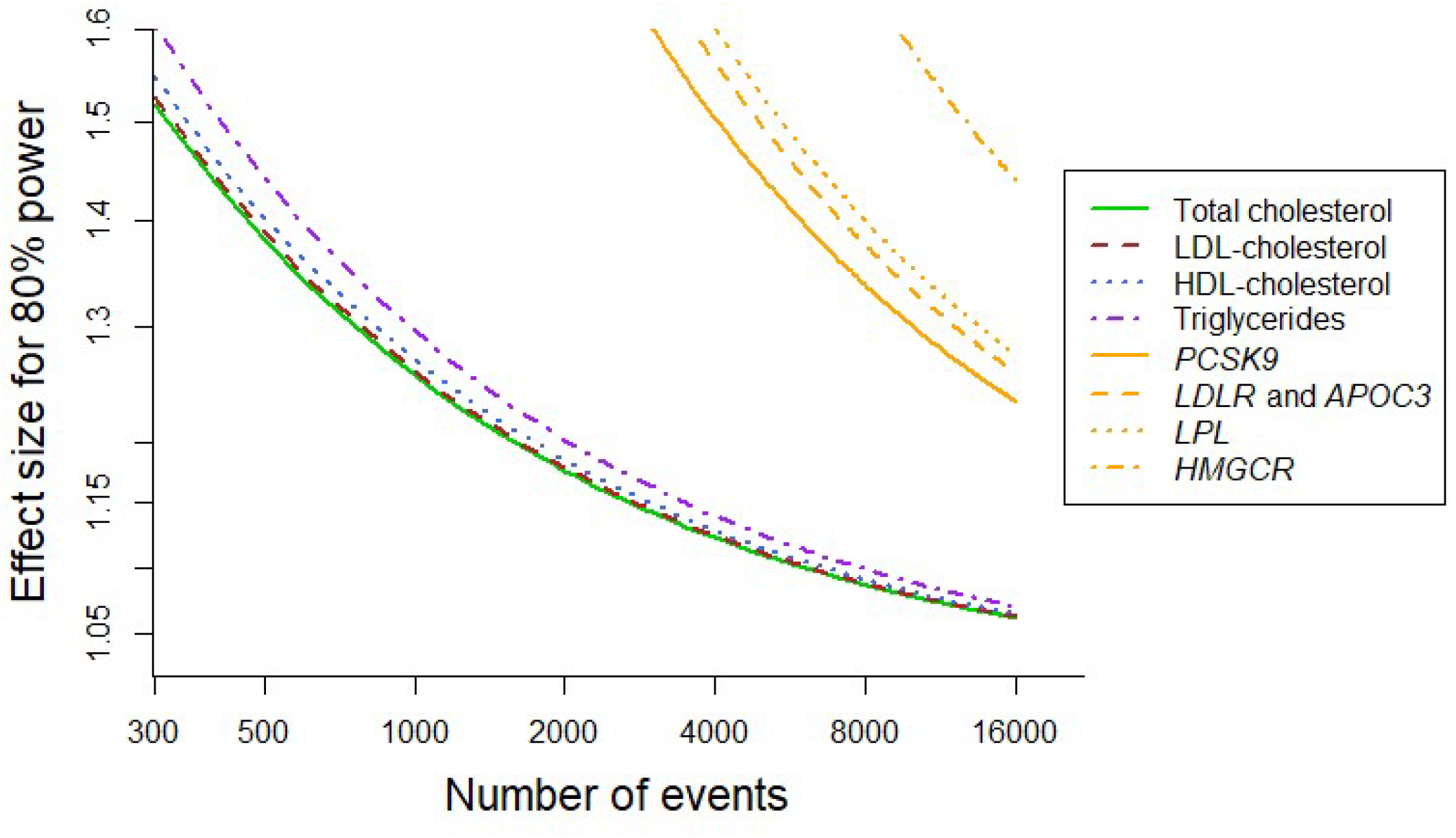
Power calculations for polygenic and gene-specific analyses, displaying the effect that can be detected with 80% power assuming a sample size of 367,703 individuals for site-specific cancers

**Supplementary Table 1:**
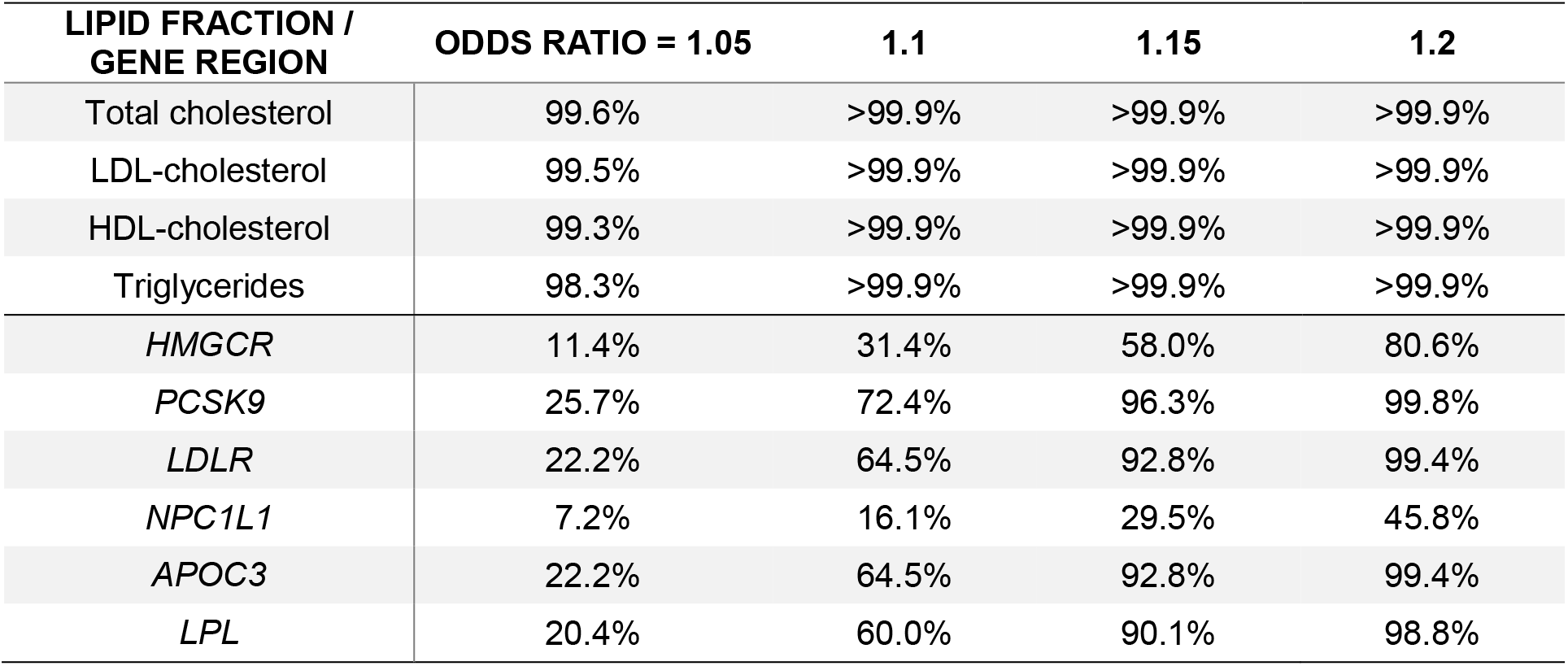
Power calculations for polygenic and gene-specific analyses, representing the power to detect a given effect size (odds ratio per 1 standard deviation increase in lipid fraction) at a significance threshold of p<0.05 for overall cancer (367,703 total individuals, 75,037 cases)

| **LIPID FRACTION /**<br>**GENE REGION** | **ODDS RATIO = 1.05** | **1.1** | **1.15** | **1.2** |
| --- | --- | --- | --- | --- |
| Total cholesterol | 99.6% | >99.9% | >99.9% | >99.9% |
| LDL-cholesterol | 99.5% | >99.9% | >99.9% | >99.9% |
| HDL-cholesterol | 99.3% | >99.9% | >99.9% | >99.9% |
| Triglycerides | 98.3% | >99.9% | >99.9% | >99.9% |
| *HMGCR* | 11.4% | 31.4% | 58.0% | 80.6% |
| *PCSK9* | 25.7% | 72.4% | 96.3% | 99.8% |
| *LDLR* | 22.2% | 64.5% | 92.8% | 99.4% |
| *NPC1L1* | 7.2% | 16.1% | 29.5% | 45.8% |
| *APOC3* | 22.2% | 64.5% | 92.8% | 99.4% |
| *LPL* | 20.4% | 60.0% | 90.1% | 98.8% |

**Supplementary Figure 2:**
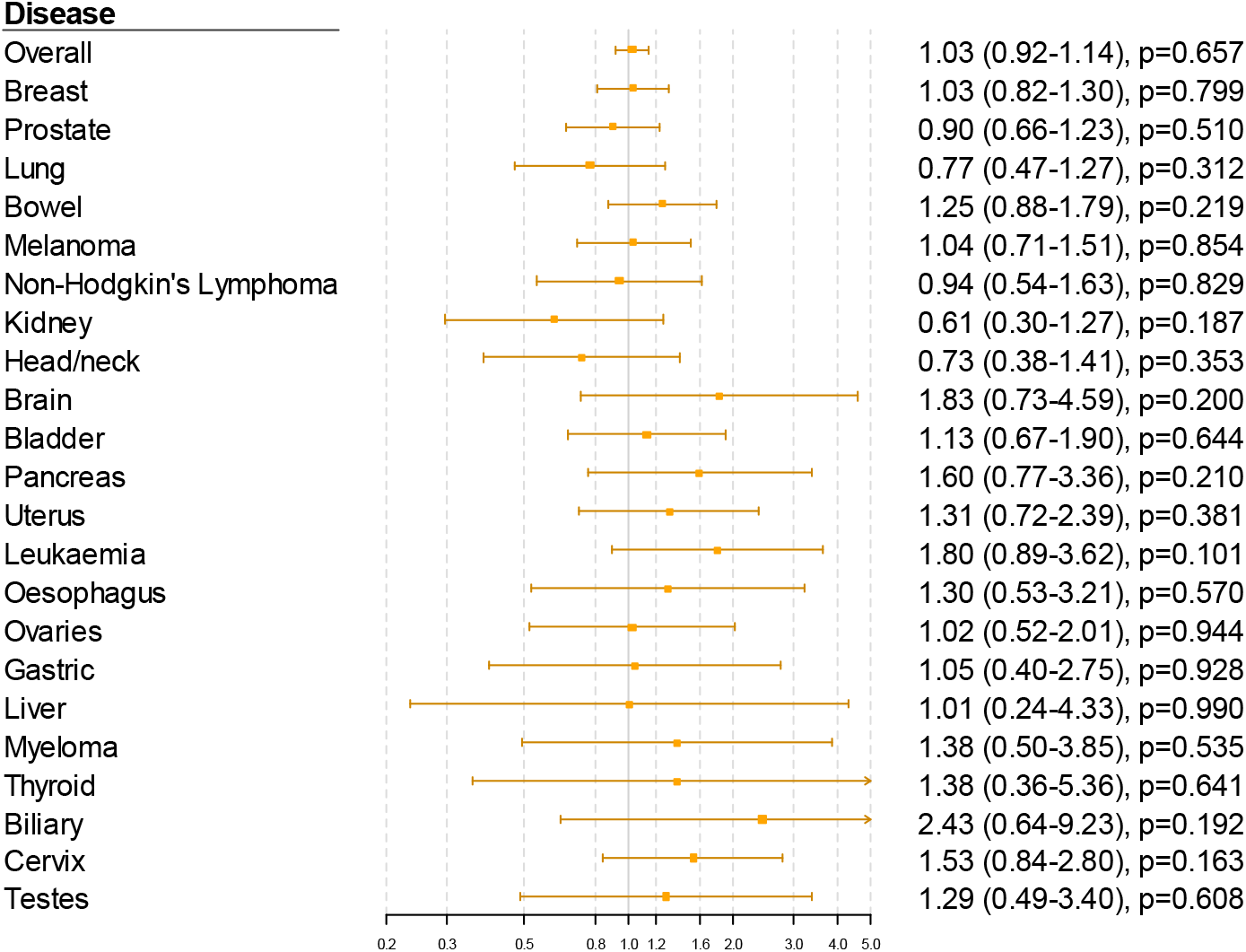
Gene-specific Mendelian randomization estimates (odds ratio with 95% confidence interval per 1 standard deviation increase in LDL-cholesterol) for variants in the *PCSK9* gene region

**Supplementary Figure 3:**
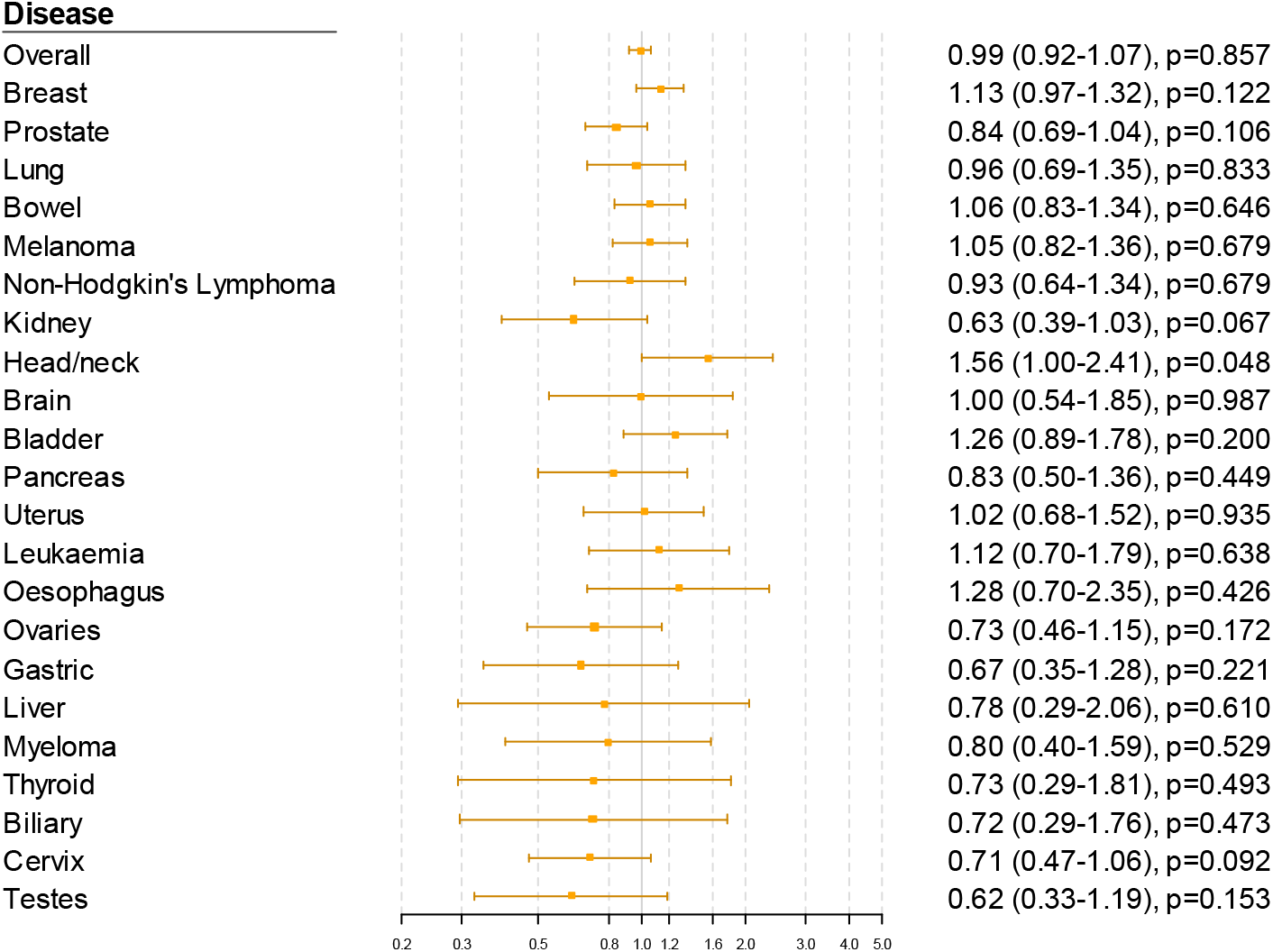
Gene-specific Mendelian randomization estimates (odds ratio with 95% confidence interval per 1 standard deviation increase in LDL-cholesterol) for variants in the *LDLR* gene region

**Supplementary Figure 4:**
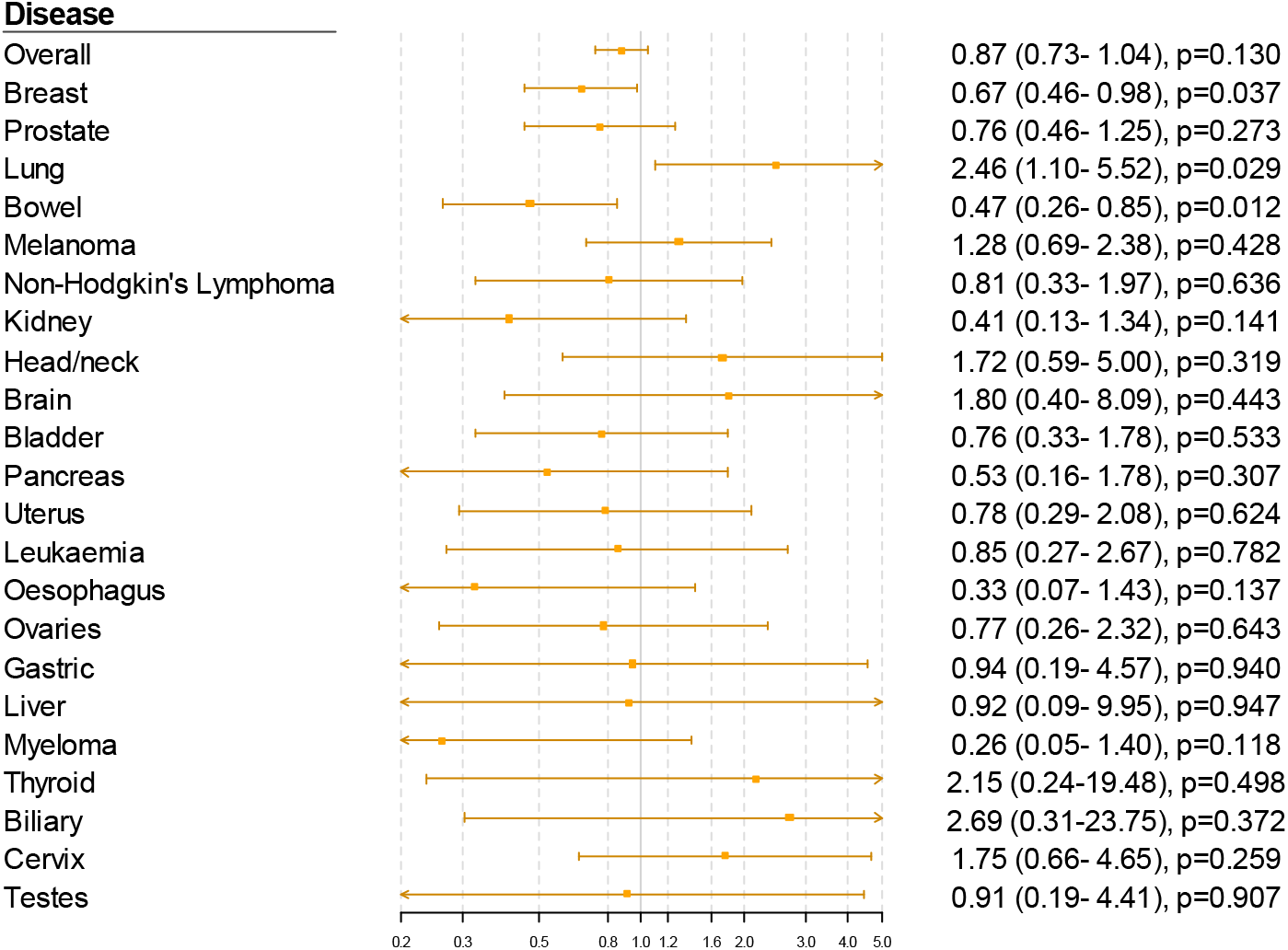
Gene-specific Mendelian randomization estimates (odds ratio with 95% confidence interval per 1 standard deviation increase in LDL-cholesterol) for variants in the *NPC1L1* gene region

**Supplementary Figure 5:**
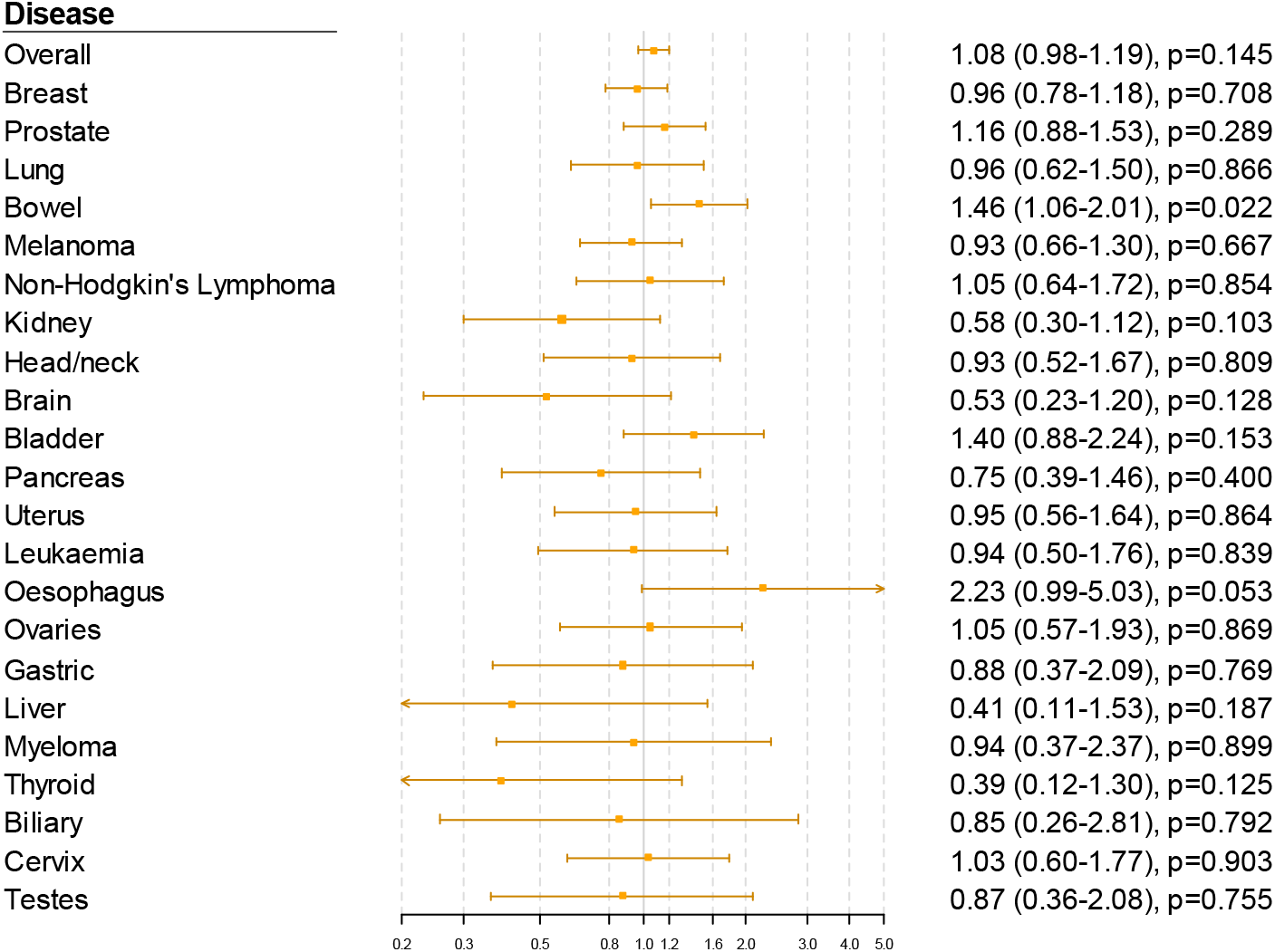
Gene-specific Mendelian randomization estimates (odds ratio with 95% confidence interval per 1 standard deviation increase in triglycerides) for variants in the *APOC3* gene region

**Supplementary Figure 6:**
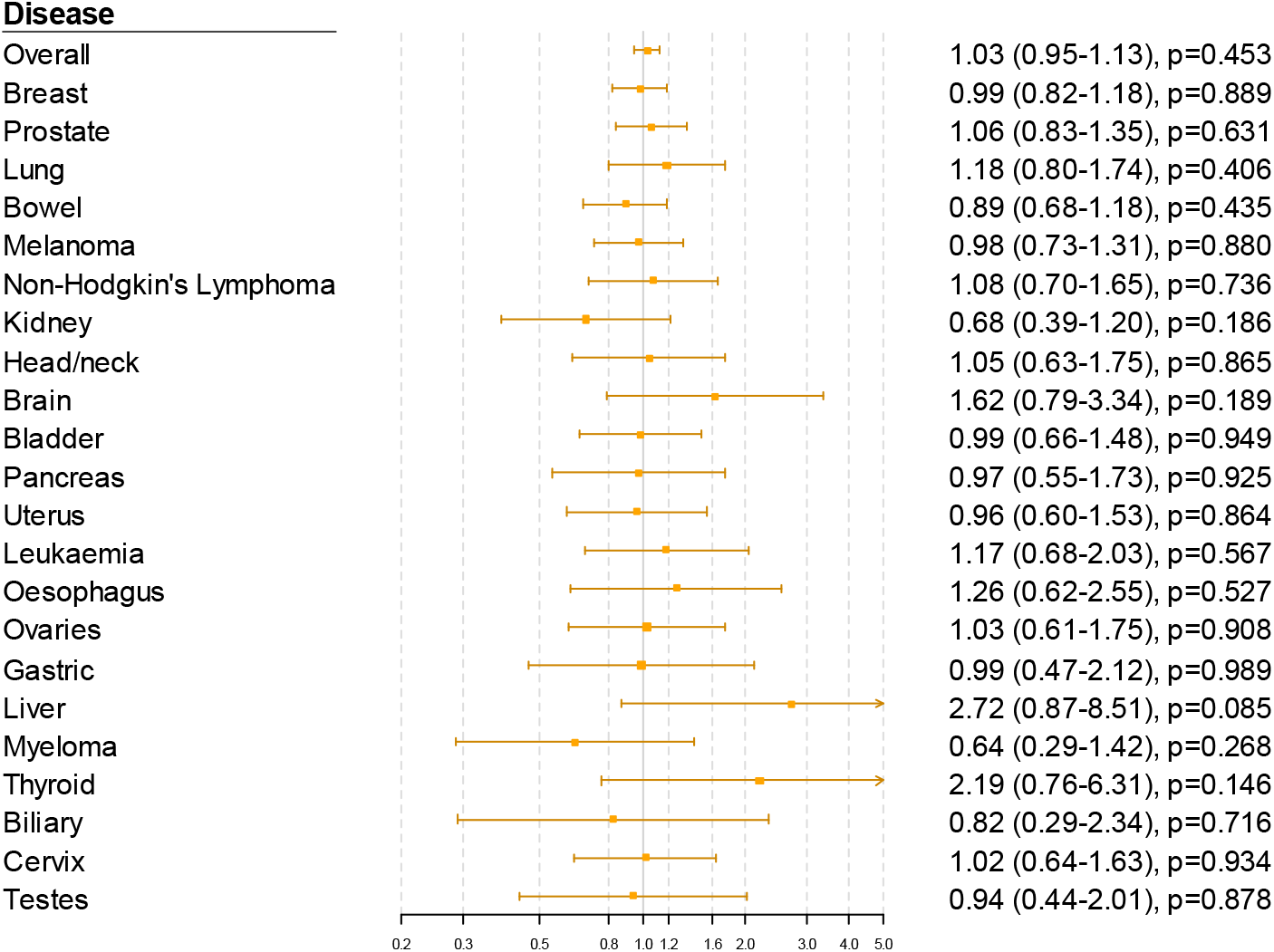
Gene-specific Mendelian randomization estimates (odds ratio with 95% confidence interval per 1 standard deviation increase in LDL-cholesterol) for variants in the *LPL* gene region

**Supplementary Figure 7:**
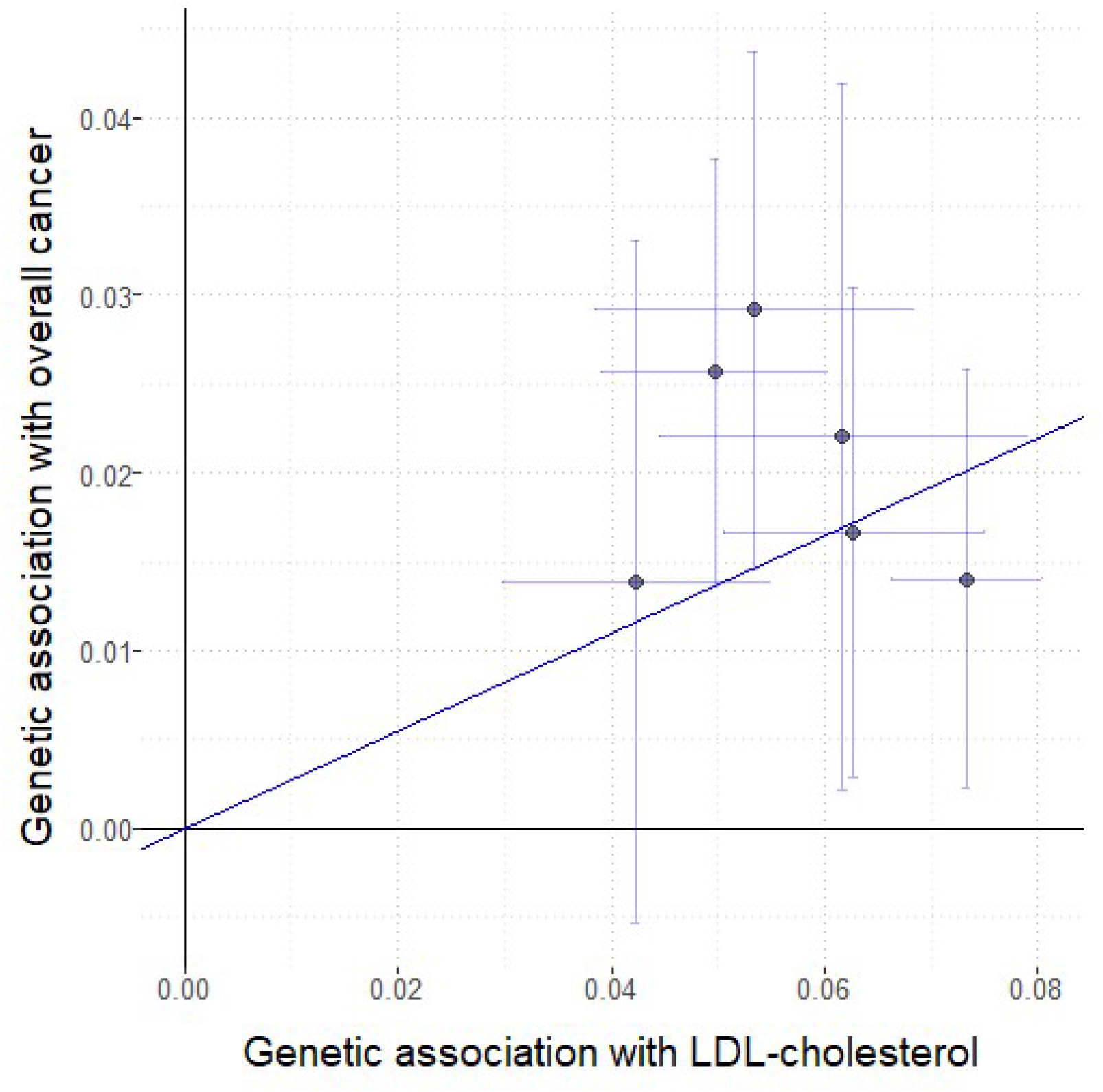
Genetic associations with LDL-cholesterol (standard deviation units) plotted against genetic associations with overall cancer (log odds ratios) for six variants in the *HMGCR* gene region

**Supplementary Table 2:** Estimates (odds ratio per 1 standard deviation increase in lipid fraction and 95% confidence interval) from polygenic multivariable Mendelian randomization analyses including all lipid-related variants. Estimates with p < 0.05 are reported in **bold**.

| CANCER SITE / CANCER TYPE | LDL-CHOLESTEROL | HDL-CHOLESTEROL | TRIGLYCERIDES |
| --- | --- | --- | --- |
| Overall cancer | 1.03 (0.99-1.07) | 1.02 (0.97-1.06) | 1.02 (0.96-1.07) |
| Breast | 1.06 (0.98-1.14) | 1.06 (0.97-1.16) | 0.96 (0.87-1.07) |
| Prostate | 0.98 (0.89-1.08) | 1.03 (0.90-1.19) | 1.03 (0.90-1.19) |
| Lung | 1.05 (0.92-1.21) | 0.95 (0.81-1.12) | 0.84 (0.69-1.01) |
| Bowel | **1.16 (1.04-1.29)** | 1.08 (0.96-1.22) | 1.02 (0.87-1.18) |
| Melanoma | 1.04 (0.93-1.15) | 0.94 (0.83-1.06) | **1.18 (1.01-1.37)** |
| Non-Hodgkin's lymphoma | 1.05 (0.91-1.23) | 1.06 (0.89-1.27) | 1.02 (0.82-1.27) |
| Kidney | 1.01 (0.83-1.23) | 0.94 (0.75-1.18) | 0.98 (0.74-1.29) |
| Head/Neck | 1.03 (0.88-1.21) | 1.03 (0.85-1.24) | 0.98 (0.78-1.24) |
| Brain | 0.98 (0.78-1.23) | 0.81 (0.62-1.07) | 0.85 (0.61-1.18) |
| Bladder | 0.99 (0.87-1.13) | 1.04 (0.89-1.22) | 1.16 (0.96-1.40) |
| Pancreas | 0.92 (0.76-1.10) | 0.89 (0.72-1.11) | 1.00 (0.77-1.30) |
| Uterus | 1.00 (0.85-1.16) | 0.96 (0.80-1.16) | 0.97 (0.78-1.21) |
| Leukaemia | 1.01 (0.85-1.20) | 1.11 (0.90-1.36) | 1.10 (0.86-1.41) |
| Oesophagus | 0.97 (0.77-1.23) | 1.08 (0.82-1.43) | 1.09 (0.78-1.53) |
| Ovaries | 1.07 (0.90-1.27) | 0.98 (0.80-1.20) | 1.09 (0.85-1.39) |
| Gastric | **0.76 (0.59-0.97)** | 1.17 (0.88-1.57) | 1.02 (0.72-1.45) |
| Liver | 0.67 (0.45-1.02) | 1.36 (0.83-2.22) | 1.12 (0.62-2.03) |
| Myeloma | 1.22 (0.93-1.59) | 1.31 (0.95-1.79) | 0.88 (0.60-1.29) |
| Thyroid | 0.89 (0.64-1.25) | 1.04 (0.70-1.55) | 0.96 (0.59-1.56) |
| Biliary | 0.86 (0.62-1.20) | 0.86 (0.58-1.28) | 0.75 (0.47-1.21) |
| Cervix | 0.97 (0.82-1.15) | 1.07 (0.88-1.30) | 1.09 (0.86-1.38) |
| Testes | 0.87 (0.68-1.11) | 0.99 (0.74-1.32) | 1.19 (0.84-1.70) |

**Supplementary Table 3:** Estimates from different Mendelian randomization methods for association between bowel cancer and genetically-predicted total cholesterol and LDL-cholesterol

| ***Bowel cancer*** | **Multivariable**<br>**IVW method** | **Univariable**<br>**IVW method** | **MR-Egger**<br>**method** | **Weighted median**<br>**method** |
| --- | --- | --- | --- | --- |
| ***Total cholesterol*** | - | 1.18<br>(1.06-1.32) | 1.10<br>(0.94-1.27) | 1.05<br>(0.91-1.22) |
| ***LDL-cholesterol*** | 1.16<br>(1.04-1.29) | 1.03<br>(0.88-1.19) | 1.09<br>(0.85-1.41) | 1.04<br>(0.86-1.26) |

**Supplementary Table 4:** Definitions of site-specific cancer outcomes in UK Biobank

| **Cancer site / type** | **ICD-9 codes** | **ICD-10 codes** | **Self-report (field 20001)** | **Cancer histology (field 40011)** |
| --- | --- | --- | --- | --- |
| *Breast* | 174., 175., V10.3 | C50., Z85.3 | 1002 |  |
| *Prostate* | 185., V10.46 | C61., Z85.46 | 1044 |  |
| *Lung* | 162., V10.1 | C33., C34., C39.9, Z85.1 | 1001, 1027, 1028, 1080 |  |
| *Bowel* | 153., 154.0, 154.1, V10.05, V10.06 | C18., C19., C20., Z85.038, Z85.048 | 1020, 1022, 1023 |  |
| *Melanoma* | 172., V10.82 | C43., Z85.820 | 1059 |  |
| *Non-Hodgkin's lymphoma* | 200., 202.0, 202.1, 202.2, 202.7, V10.71 | C82., C83., C84., C85., C86., C88.0, C88.4, Z85.72 | 1053 |  |
| *Kidney* | 189.0, V10.52 | C64., Z85.528 | 1034 |  |
| *Head/Neck* | 140., 141., 142., 143., 144., 145., 146., 147., 148., 149., 160., 161., V10.01, V10.02, V10.21, V10.22 | C00., C01., C02., C03., C04., C05., C06., C07., C08., C09., C10., C11., C12., C13., C14., C30., C31., C32., Z85.21, Z85.22, Z85.81 | 1006, 1007, 1009, 1004, 1010, 1011, 1012, 1077, 1078, 1079, 1005, 1015, 1016 |  |
| *Brain* | 191., 192.0, 192.1, 192.2, 192.3, V10.85 | C70., C71., C72.0, C72.3, Z85.841 | 1031, 1032, 1033 |  |
| *Bladder* | 188., 189.1, 189.2, V10.51, V10.53 | C67., C65., C66., Z85.51, Z85.54, Z85.53 | 1035 |  |
| *Pancreas* | 157. | C25., Z85.07 | 1034 |  |
| *Uterus* | 179., 182., V10.42, | C54., C55., Z85.42 | 1040 |  |
| *Leukaemia* | 204., 205., 206., 207., 208., V10.6 | C91., C92., C93., C94.0, C94.2, C94.3, C94.4, C94.8, C95, Z85.6 | 1048, 1055, 1056, 1074 |  |
| *Oesophagus* | 150., V10.03 | C15., Z85.01 | 1017 |  |
| *Ovaries* | 183.0, 183.2, 183.8, 183.9, V10.43 | C56., C57.0, C57.4, Z85.43 | 1039 |  |
| *Gastric* | 151., V10.04 | C16., Z85.028 | 1018 |  |
| *Liver* | 155.0 | C22.0 | 1024 | 8170, 8171, 8172, 8173, 8174, 8175 |
| *Myeloma* | 203.0, 203.1, | C90.0, C90.1 | 1050 | 9732, 9733 |
| *Thyroid* | 193., V10.87 | C73., Z85.850 | 1065 |  |
| *Biliary* | 155.1, 156.0 | C22.1, C23., C24. | 1025 |  |
| *Cervix* | 180., V10.41 | C53., Z85.41 | 1041 |  |
| *Testes* | 186., V10.47 | C62., Z85.47 | 1045 |  |

**Supplementary Table 5:** Genetic variants included in gene-specific analyses for each region

| GENE<br>REGION | SNP | POS (HG19) | A1 | A2 | EAF | EXPOSURE |  |  | OUTCOME |  |  |
| --- | --- | --- | --- | --- | --- | --- | --- | --- | --- | --- | --- |
|  |  |  |  |  |  | BETA | SE | P | BETA | SE | P |
| *HMGCR* | rs2006760 | chr5:74562029 | G | C | 0.229 | 0.053 | 0.008 | 1.7x10<sup>-13</sup> | 0.029 | 0.007 | 8x10<sup>-5</sup> |
| *HMGCR* | rs2303152 | chr5:74641707 | G | A | 0.890 | -0.042 | 0.006 | 1.1x10<sup>-9</sup> | -0.014 | 0.010 | 0.158 |
| *HMGCR* | rs17238484 | chr5:74648496 | G | T | 0.763 | -0.063 | 0.006 | 1.4x10<sup>-21</sup> | -0.017 | 0.007 | 0.018 |
| *HMGCR* | rs12916 | chr5:74656539 | C | T | 0.412 | 0.073 | 0.004 | 7.8x10<sup>-78</sup> | 0.014 | 0.006 | 0.020 |
| *HMGCR* | rs10066707 | chr5:74560579 | A | G | 0.396 | 0.050 | 0.005 | 3.0x10<sup>-19</sup> | 0.026 | 0.006 | 3x10<sup>-5</sup> |
| *HMGCR* | rs5909 | chr5:74656175 | A | G | 0.100 | 0.062 | 0.009 | 5.0x10<sup>-13</sup> | 0.022 | 0.010 | 0.030 |
| *PCSK9* | rs2479394 | chr1:55486064 | G | A | 0.281 | 0.039 | 0.004 | 1.6x10<sup>-19</sup> | 0.001 | 0.007 | 0.892 |
| *PCSK9* | rs11206510 | chr1:55496039 | C | T | 0.172 | -0.083 | 0.005 | 2.4x10<sup>-53</sup> | 0.002 | 0.008 | 0.764 |
| *PCSK9* | rs2149041 | chr1:55502137 | C | G | 0.823 | -0.064 | 0.005 | 1.4x10<sup>-35</sup> | 0.005 | 0.008 | 0.474 |
| *PCSK9* | rs10888897 | chr1:55513061 | C | T | 0.680 | -0.064 | 0.004 | 2.5x10<sup>-50</sup> | -0.002 | 0.006 | 0.784 |
| *PCSK9* | rs7552841 | chr1:55518752 | C | T | 0.596 | 0.051 | 0.004 | 8.4x10<sup>-31</sup> | 0.008 | 0.006 | 0.214 |
| *PCSK9* | rs562556 | chr1:55524237 | A | G | 0.631 | -0.037 | 0.004 | 5.4x10<sup>-15</sup> | -0.010 | 0.008 | 0.184 |
| *LDLR* | rs6511720 | chr19:11202306 | G | T | 0.902 | 0.221 | 0.006 | 3.9x10<sup>-262</sup> | -0.002 | 0.009 | 0.810 |
| *LDLR* | rs688 | chr19:11227602 | C | T | 0.553 | -0.054 | 0.004 | 1.0x10<sup>-43</sup> | -0.001 | 0.006 | 0.913 |
| *NPC1L1* | rs10234070 | chr7:44537696 | T | C | 0.899 | 0.024 | 0.005 | 1.5x10<sup>-6</sup> | -0.010 | 0.009 | 0.280 |
| *NPC1L1* | rs2073547 | chr7:44582331 | G | A | 0.195 | 0.039 | 0.004 | 1.9x10<sup>-21</sup> | -0.007 | 0.008 | 0.326 |
| *NPC1L1* | rs217386 | chr7:44600695 | A | G | 0.586 | -0.029 | 0.003 | 1.2x10<sup>-19</sup> | 0.011 | 0.006 | 0.072 |
| *NPC1L1* | rs7791240 | chr7:44602589 | C | T | 0.084 | -0.034 | 0.005 | 1.8x10<sup>-10</sup> | -0.004 | 0.010 | 0.712 |
| *NPC1L1* | rs2300414 | chr7:44682938 | A | G | 0.069 | 0.028 | 0.006 | 5.4x10<sup>-6</sup> | -0.013 | 0.012 | 0.251 |
| *APOC3* | rs10790162 | chr11:116639104 | A | G | 0.091 | 0.231 | 0.006 | 1.1x10<sup>-249</sup> | 0.013 | 0.012 | 0.263 |
| *APOC3* | rs603446 | chr11:116654435 | C | T | 0.553 | 0.050 | 0.003 | 3.9x10<sup>-43</sup> | 0.012 | 0.006 | 0.038 |
| *LPL* | rs1801177 | chr8:19805708 | A | G | 0.011 | 0.164 | 0.023 | 1.1x10<sup>-9</sup> | -0.015 | 0.023 | 0.508 |
| *LPL* | rs268 | chr8:19813529 | A | G | 0.986 | -0.280 | 0.035 | 2.2x10<sup>-16</sup> | -0.027 | 0.022 | 0.214 |
| *LPL* | rs301 | chr8:19816934 | C | T | 0.459 | -0.109 | 0.004 | 1.9x10<sup>-167</sup> | 0.003 | 0.007 | 0.654 |
| *LPL* | rs326 | chr8:19819439 | A | G | 0.673 | 0.087 | 0.005 | 1.0x10<sup>-63</sup> | -0.006 | 0.006 | 0.334 |
| *LPL* | rs328 | chr8:19819724 | C | G | 0.870 | 0.167 | 0.006 | 2.0x10<sup>-179</sup> | 0.010 | 0.010 | 0.284 |

SNP = single nucleotide polymorphism, POS (HG19) = chromosome and genomic position, A1 = effect allele, A2 = other allele, EAF = effect allele frequency, EXPOSURE = LDL-cholesterol for variants in *HMGCR*, *PCSK9* and *LDLR* gene regions, and triglycerides for *APOC3* and *LPL* gene regions, OUTCOME = overall cancer, BETA = beta-coefficient (in standard deviation units for exposure, log odds ratio for outcome) from univariable (marginal) regression analysis, SE = standard error, P = p-value.

